# A double-blind trial of decoded neurofeedback intervention for specific phobias

**DOI:** 10.1101/2023.04.25.23289107

**Authors:** Cody A. Cushing, Hakwan Lau, Mitsuo Kawato, Michelle G. Craske, Vincent Taschereau-Dumouchel

## Abstract

**Aim:** A new closed-loop fMRI method called multi-voxel neuro-reinforcement has the potential to alleviate the subjective aversiveness of exposure-based interventions by directly inducing phobic representations in the brain, outside of conscious awareness. The current study seeks to test this method as an intervention for specific phobia.

**Methods:** In a randomized, double-blind, controlled single-university trial, individuals diagnosed with at least two (1 target, 1 control) animal subtype specific phobias were randomly assigned (1:1:1) to receive 1, 3, or 5 sessions of multi-voxel neuro-reinforcement in which they were rewarded for implicit activation of a target animal representation. Amygdala response to phobic stimuli was assessed by study staff blind to target and control animal assignments. Pre-treatment to post-treatment differences were analyzed with a 2-way repeated-measures ANOVA.

**Results:** A total of 23 participants (69.6% female) were randomized to receive 1 (n=8), 3 (n=7), or 5 (n=7) sessions of multi-voxel neuro-reinforcement. Eighteen (n=6 each group) participants were analyzed for our primary outcome. After neuro-reinforcement, we observed an interaction indicating a significant decrease in amygdala response for the target phobia but not the control phobia. No adverse events or dropouts were reported as a result of the intervention.

**Conclusion:** Results suggest multi-voxel neuro-reinforcement can specifically reduce threat signatures in specific phobia. Consequently, this intervention may complement conventional psychotherapy approaches with a non-distressing experience for patients seeking treatment. This trial sets the stage for a larger randomized clinical trial to replicate these results and examine the effects on real-life exposure.

**Clinical Trial Registration:** The now-closed trial was prospectively registered at ClinicalTrials.gov with ID NCT03655262.

## Introduction

Fear-based disorders such as specific phobia are among the most difficult mental disorders to treat. The most widely empirically supported treatment is ‘exposure therapy’, which involves direct exposure to fear-causing or panic-inducing stimuli (1). This treatment is highly effective in reducing fear. However, conscious exposure to feared stimuli is a disturbing and unpleasant experience for the patient, leading to high rates of attrition (2,3). Exposure treatment dropout rates can be as high as 70%, with 60% being unwilling to even start treatment (1,3–5). As a result, only a small percentage of patients can actually benefit from an otherwise effective treatment.

Consequently, neurofeedback has been explored as a way of directly regulating brain activity in a number of mental health disorders (6–12). A promising new fMRI method called multi-voxel neuro-reinforcement (13–15) has demonstrated the ability to lessen physiological defensive responses to both laboratory-conditioned fears and pre-existing fears through a kind of ‘unconscious exposure’ (16–19). Exposure treatments bypassing conscious awareness have shown promise in reducing fear responses in phobic individuals (20) owing to the dissociability of threat response and learning from subjective experience (21,22). By using a machine-learning classifier (also referred to as a ‘decoder’), neuro-reinforcement can be provided based on a specific stimulus category (e.g. spider) rather than average brain activity alone (19). Critically, this results in no subjective discomfort for the patient, but yet can still lead to lasting reduction of fear (7,8,18,23–25).

A decoder can be built for a patient with a phobia using brain data from a group of healthy controls for whom viewing repeated images of a target representation (e.g. spider) produces no fear reaction (Fig. 1). Training between-subject decoders this way enables “nonconscious exposure” in patients with phobias, without exposing them to feared stimuli. This surrogate data approach was explored in our previous proof-of-concept study (17), but there participants still saw the feared images during the decoder construction task. In the present study, we test for the first time whether multi-voxel neuro-reinforcement can succeed using a decoder trained completely on surrogate data where the participant undergoing neuro-reinforcement has never seen the feared images.

**Figure 1.**
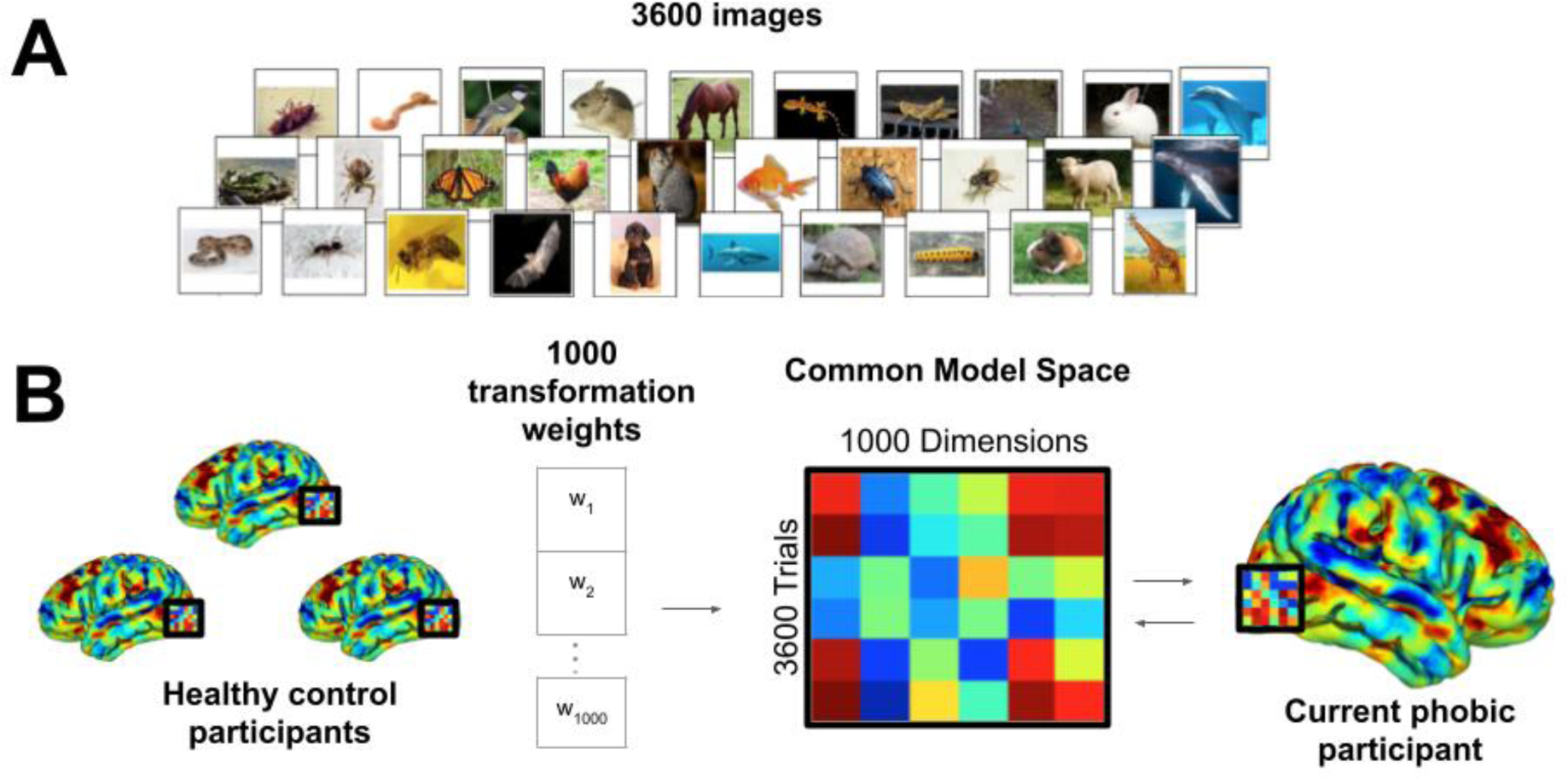
Functional alignment of brain data into phobic participant brain using hyperalignment. (A) All participants complete a near-identical task in the fMRI scanner where 3600 images are rapidly viewed during 0.98 second presentations. Phobic patients view happy human faces instead of their own phobic categories. Healthy controls view images from all categories. (B) Transformation parameters into the functionally aligned common model space are determined with phobic image trials withheld. Data from all participants for all categories (including phobic categories) are transformed into the common model space and then reverse transformed into the native space of the current phobic participant. A machine-learning classifier can then be trained on phobic images in the phobic participant’s native brain space despite the participant never having personally viewed the images.

Here, we describe a preregistered (Phase 1: https://osf.io/gjvmt?view_only=b6827aa394f143aeb29b99c095bd4183) double-blind placebo-controlled clinical trial of this method as an intervention in a population with specific phobia. We preregistered 5 hypotheses (H1-5). We hypothesized that amygdala responses (H1) and skin conductance responses (H2) to phobic stimuli would selectively decrease for the targeted phobia relative to the control phobia following neuro-reinforcement. We focus on amygdala responding as our primary outcome due to its canonical role in learning and extinction of threat and fear responses (26–30). Additionally, we hypothesized that subjective fear ratings would stay the same following neuro-reinforcement (H3), despite the predicted changes in physiological responses, based on our previous findings in a non-clinical population (17).

Secondarily, we introduce a modified affective Stroop task in which participants make rapid size judgments about phobic and neutral stimuli. In this task, we hypothesized that reaction times would be slower for phobic stimuli (H4i) and that following neuro-reinforcement there would be a selective reduction in reaction times (H4ii) and amygdala responses (H4iii) in response to the targeted phobia category compared to the control phobia. Finally, we randomly assigned participants to receive either 1, 3, or 5 sessions of neuro-reinforcement. We hypothesized that those receiving the most neuro-reinforcement would demonstrate the largest effects (H5).

To anticipate, we did not manage to collect the full amount of data (N=30) as planned, due to pandemic-related circumstances. However, despite the reduced sample size (N=18), our primary hypothesis about amygdala response reduction (H1) was confirmed. Additionally, we found mixed evidence in support of secondary hypotheses concerning attentional capture by phobias in our novel affective Stroop task (H4). Unfortunately, due to circumstances outside our control, we lacked the statistical power to adequately assess the between-group differences for the amount of neuro-reinforcement received (H5).

## Methods

### III. Trial Design and Participant Screening

The current trial was designed as a randomized, within-subject controlled, experimenter and participant blinded dose-response study with randomization to 1, 3, or 5 days of multi-voxel neuro-reinforcement and a primary endpoint of amygdala activation to targeted phobic stimuli compared to control phobic stimuli. The study protocol was approved by the Institutional Review Board at the University of California, Los Angeles concordant with the provisions of the Declaration of Helsinki. Specific phobias were diagnosed using the Anxiety Disorders Interview Schedule-5 (31) in a diagnostic interview conducted by trained and reliability certified study staff. Details of diagnostic screening and control vs phobia grouping can be found in *Supplemental Methods*.

For multi-voxel neuro-reinforcement, 23 participants (mean age (s.d.) = 26.5 (9.40), 69.6% female) with at least two specific animal phobias were enrolled for treatment. The informed consent of participants was obtained pursuant to the procedures of the Institutional Review Board at the University of California, Los Angeles. Participants were randomly assigned to complete either 1 (N=8), 3 (N=7), or 5 (N=8) days of multi-voxel neuro-reinforcement to determine the dose-response relationship with clinical outcomes. Of these 23 participants, 2 did not finish multi-voxel neuro-reinforcement (1 due to technical issues, 1 due to scheduling issues). The PI (MGC) monitored the study on a day-to-day basis with prompt reporting of adverse events to the IRB, NIMH, and other agencies as appropriate. The following adverse events were monitored: deaths, suicide attempts, study dropout, psychiatric hospitalizations, and clinical deterioration as defined as emergent suicidal ideation or suicidal plan, development of serious substance abuse, or the emergence of a new psychiatric or medical diagnosis or behavior posing a significant risk to the subjects of others. Zero adverse events were recorded. Outcome analyses were performed on participants that completed the clinical trial per protocol. Of the 21 participants who completed multi-voxel neuro-reinforcement, 1 experienced nausea during tasks and was excluded from further analysis. Two participants did not complete the pre-post “fear test” task properly (closed their eyes or turned away in response to phobic images) for amygdala response (described below) and were excluded from analyses relevant to that task, leaving 18 participants (n=6 each dosage group) for our primary analyses (H1, H2, H3, and H5). This cohort of 18 participants falls short of our original goal of 30 participants because our funding expired due to the shutdowns and recruitment difficulties resulting from the COVID-19 global pandemic. As a result, any further data collection was impossible. For secondary analysis of the affective Stroop task (H4), 2 of the 18 participants included in the fear test analysis did not complete the affective Stroop task, and one participant that did not complete the fear test task properly, but did complete the affective Stroop task, resulting in 17 participants analyzed (H4).

### II. Randomization and masking

Upon enrollment, participants were randomized to either 1, 3, or 5 sessions of neuro-reinforcement using a random number generator by study coordinators with a 1:1:1 allocation. This randomization was not directly investigated during our primary analyses due to COVID-19 restrictions on data collection that limited power to detect between-group differences. However, randomization group was controlled for as a covariate in all primary analyses. Neuro-reinforcement itself was then controlled with a double-blind within-subject placebo. Participants had at least 2 phobias with one phobia being used as the treatment target while another served as control. Assignment of the target and control phobias was performed automatically by a computer during data processing according to the procedures outlined in *Supplemental Methods*.

Experimenters were blinded during data collection by loading the target pattern automatically with computer software. Target and control pattern labels were automatically saved during decoder construction processing (outlined below) and stored as MATLAB variables to be automatically loaded during post-treatment offline analysis on computers not involved in data collection. Participants were blinded to the target of their treatment as neuro-reinforcement was performed implicitly; i.e., participants were provided no specific instruction as to what to think about during neuro-reinforcement and had no knowledge as to which phobia was being targeted, or how many of their multiple phobias would be targeted. Participant strategies were monitored daily to ensure participants had not coincidentally thought about their target (or control) phobia during neuro-reinforcement, effectively unblinding themselves. No participants reported thinking about either the target or control phobia during neuro-reinforcement.

### III. Decoder Construction

Prior to neuro-reinforcement, a between-subject machine-learning decoder was trained for the target phobic image category in the ventral temporal (VT) area (Fig. 1). The decoder was constructed using brain data from healthy controls (N=22) using a functional alignment method called hyperalignment (32). During an initial fMRI session (Fig. 2A), each healthy control viewed the same image dataset of 3600 images consisting of 40 categories of animals and objects (e.g. birds, butterflies, snakes, spiders) (Fig. 1A). Conversely, participants with phobias viewed the same image dataset but with their specific phobias removed to avoid unnecessary exposure.

**Figure 2.**
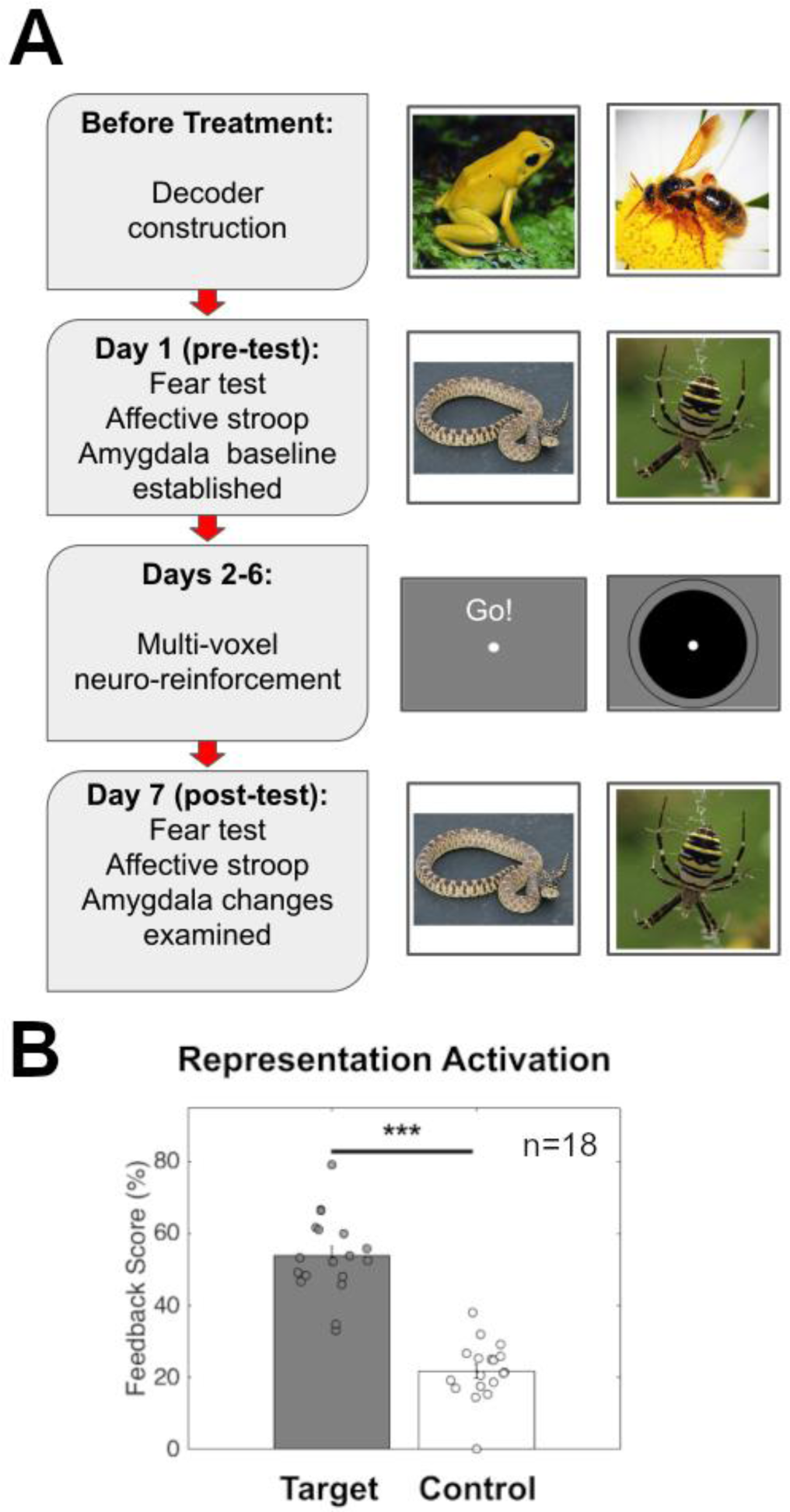
Study design and activation of Target and Control representations. (A) Timeline detailing patient activities during each day’s fMRI session with sample stimuli from each day. Before beginning the treatment program patients undergo a decoder construction session where they view non-phobic images to enable hyperalignment with healthy control subjects. On day 1 of treatment, patients complete a pre-test in which phobic (and non-phobic) images are rated for fearfulness. Over the next 5 days, patients complete their assigned number of multi-voxel neuro-reinforcement sessions (1, 3, or 5 days). On day 7, patients complete the same task as a post-test to assess changes in amygdala and SCR response to treated and untreated phobias. (B) Representation pattern activation (measured by feedback score) for Target phobia compared to Control phobia. Target phobia pattern was activated significantly more than Control during neuro-reinforcement. *** *p*<0.001

Participant-specific decoders were developed using surrogate data based on previous methods (17), detailed in *Supplemental Methods* along with task details (Fig. 1B). Importantly, the VT area is a category-selective visual region in which hyperalignment decoding is not affected by fear levels, enabling the use of surrogate brain data from healthy controls to train decoders for phobic participants (17). Additional research has shown that some ventral visual areas can appear to be predictive of subjective fear ratings (21). However, these findings likely do not truly reflect “subjective fear” but rather statistical regularities in commonly feared stimuli. This appears to be the case since the same “fear-brain” associations of a fearful person can be detected when using brain data from persons reporting no fear of the stimuli. These findings suggest that this brain signal does not truly represent fear but perhaps statistical regularities of commonly feared animal categories (e.g. spiders). This further suggests that VT representations should be similar across participants regardless of fear levels. Of the potential phobic categories to be selected for treatment for the current participant, the phobia with the highest cross-validated AUC scores was blindly selected via computer program as the target for treatment. The within-subject control was also blindly selected through automated random selection from the remaining phobic categories if the participant had more than two phobias. Double-blind target selection was performed in this manner to maximize signal to noise during neuro-reinforcement. Importantly, decoder AUC during decoder construction should not influence the value of feedback scores during neuro-reinforcement. Confirming this, in this study there was no relation between decoder performance during decoder construction and neuro-reinforcement scores for the target category (r(16)=-0.11, p=0.66, Supplemental Figure S1A). Moreover, concerning the difference between target and control categories, there was no relation between the difference in decoder performance during decoder construction and the difference in scores calculated during neuro-reinforcement (r(15)=-0.096, p=0.71, Supplemental Figure S1B).

### IV. Pre- and Post-Neuro-Reinforcement Assessments

Each phobic participant completed a pre-treatment and post-treatment fMRI session (Fig. 2A), during which they completed a fear test as well as an affective Stroop task while their BOLD activity was recorded.

*Fear test (H1, H2, H3, and H5*). Neural and subjective fear responses were measured to 6-second exposures to photographic images from phobic and neutral animal categories, that included the targeted and control phobias for each participant, following the previous proof-of-concept study (17). The neutral animal category was randomly selected from animals for which participants self-reported a total absence of fear at screening. Participants reported how fearful each image made them feel on a 7-point likert scale following each image presentation. See *Supplemental Methods* for full task details.

*Skin Conductance Response (H2).* Skin Conductance Response (SCR) recordings were taken in the fMRI scanner during the fear test. Details of data collection and analysis are reported in *Supplemental Methods*.

*Affective Stroop (H4).* An affective Stroop task assessed reflexive attentional responses to phobic stimuli. Participants made rapid judgments about whether briefly presented animals could fit in their hand. Full task details can be found in *Supplemental Methods*.

### V. Multi-voxel neuro-reinforcement

Using multi-voxel neuro-reinforcement, successful activation of the phobic image category was paired with reward (Fig. 2A). Participants were only aware that the neuro-reinforcement task was intended to function as a treatment for phobia. They were blinded to all other information including what the feedback was based on in their brain, how it was calculated, or how many of their phobias this would treat. While participants laid in the fMRI scanner instructed to “use whatever mental strategy they can” to get the best feedback, a neuro-reinforcement method (17) was used to reward a nonconsciously represented phobic image category (e.g., spider).

Feedback was based on real-time output of the decoder constructed for the individual corresponding to the specific animal phobia selected for targeting. Individual strategies were recorded at the end of each multi-voxel neuro-reinforcement session. Common strategies reported by participants included thinking about family and friends, memories from the past or plans for the future, imagining oneself doing activities (exercising, sports, riding rollercoasters, etc.), or “mindful” techniques like focusing on the breath, doing mental math, or simply trying to imagine the feedback circle getting bigger. Participants reported that strategies that seemed to work at one moment did not seem to work later, indicating that one strategy did not work better than others overall.

Each neuro-reinforcement run began with an extended rest period of 50 seconds. Then, an additional rest period of 10 seconds was collected to determine baseline BOLD activity levels followed by 16 trials of neuro-reinforcement. Each trial began with 6 seconds of rest, followed by 6 seconds of “induction” where participants modulated their brain activity in an attempt to receive high feedback. Following induction, real-time decoder output was calculated during a 4-second period and then displayed as a green disc for 2 seconds. This calculation focused on BOLD activity from 4 seconds after the start of the induction period until 4 seconds after the induction period ended to account for hemodynamic response delay. The size of the disc directly corresponded to the likelihood estimate such that a 100% likelihood was associated with a maximum disc size (indicated by a visual boundary) and a 0% likelihood was associated with no disc display.

### VI. Data Processing

*Amygdala Response Analysis (H1, H4iii and H5) and Affective Stroop (H4). See Supplemental Methods*.

### VII. Data Analysis Plan

Amygdala responses were tested with a 2 (condition: target phobia/control phobia) x 2 (time: pre-treatment/post-treatment) repeated-measures ANOVA using JASP software (JASP Team 2022). Due to limited sample size (from the COVID-19 pandemic), we were insufficiently powered to analyze neuro-reinforcement dosage groups separately, as we had initially preregistered in hypothesis H5. Instead, neuro-reinforcement group (1, 3, or 5 days) was included as a covariate in the ANOVA as was each participant’s total number of phobias, as a measure of clinical severity. The between-group data are presented in *Supplemental Figures S3A, S3B, & S3C and S4A, S4B, & S4C* for illustration purposes and the preregistered statistical analysis for H5 is reported in *Supplemental Results*. To test for a significant reduction in amygdala response for the target phobia category post-treatment compared to the control phobia (H1), we used contrasts of marginal means in JASP to test effects within our ANOVAs which included covariates. These follow-up contrasts were performed on pre- and post-treatment activations for the target phobia and control phobia.

Planned t-tests were performed on pre- and post-treatment subjective fear ratings for the target phobia and control phobia, using custom scripts in Matlab, to test H3. One of the 18 participants was excluded from this analysis of self-reported fear due to not using the button box properly, resulting in 17 participants.

To verify phobic images were modulating attention as intended, a t-test was performed on affective Stroop reaction times to phobic images (grouping target and control) and neutral animal images pre-treatment (H4.i). For treatment effects (H4.ii), reaction times for correct trials were tested with a 2 (condition: target phobia/control phobia) x 2 (time: pre-treatment/post-treatment) repeated-measures ANOVA using JASP software (JASP Team 2022). Dosage group and number of phobias were included as covariates in the model. Similar to the amygdala response analysis, contrasts of marginal means were performed on pre- and post-treatment reaction times for the target phobia and control phobia.

## Results

A total of 23 participants (Table 1, Table 2, Fig. 3, Supplemental Table 1) completed pre-treatment. Our intended goal of 30 participants, 10 per neuro-reinforcement dosage condition, was not achievable due to COVID-19 difficulties.

**Figure 3.**
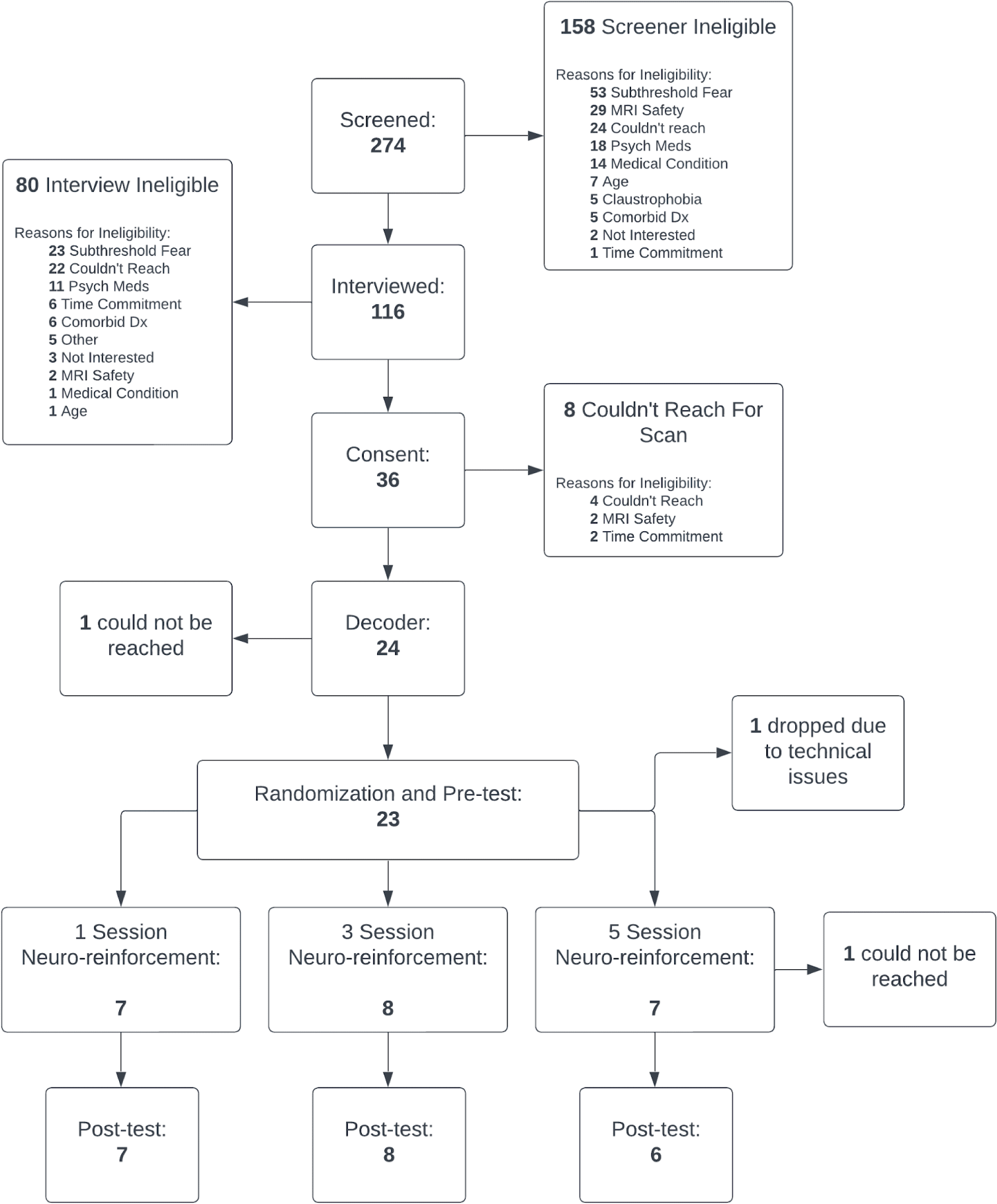
CONSORT diagram of recruitment flow.

**Table 1.** Participant demographics.

| **Race** |  |
| --- | --- |
| White | 9 |
| Black | 2 |
| Asian/Pacific Islander | 9 |
| Other | 1 |
| Not Reported | 2 |
| **Ethnicity** |  |
| Hispanic | 5 |
| Non-Hispanic | 18 |
| **Gender** |  |
| Male | 7 |
| Female | 16 |
| Non-binary | 0 |
| **Age:** mean(s.d.) | 26.5 (9.40) |
| **Education Level** |  |
| High School | 4 |
| Some College | 3 |
| Associates/2-year degree or higher | 16 |

**Table 2.** Participant measures (collapsed across randomization groups)

| **Baseline** |  |  |
| --- | --- | --- |
|  | **Target Phobia** | **Control Phobia** |
| ADIS-5 Interviewer Fear Rating (0-8): mean (s.d.) | 5.17 (1.11) | 5.91 (1.00) |
| ADIS-5 Interviewer Avoidance Rating (0-8): mean (s.d.) | 5.30 (1.43) | 5.78 (1.24) |
| **Pre-treatment** |  |  |
|  | **Target Phobia** | **Control Phobia** |
| Fear Test Amygdala: mean (s.d.) Beta value | 0.50 (0.96) | 0.31 (0.57) |
| Fear Test Subjective Fear Rating (0-8): mean (s.d.) | 3.63 (1.44) | 4.20 (1.12) |
| Affective Stroop Amygdala: mean (s.d.) Beta value | 0.01 (0.31) | 0.07 (0.28) |
| Affective Stroop Reaction Time: mean (s.d.) seconds | 1.00 (0.20) | 1.01 (0.12) |
| **Post-treatment** |  |  |
|  | **Target Phobia** | **Control Phobia** |
| Fear Test Amygdala: mean (s.d.) Beta value | -0.10 (0.64) | -0.13 (0.86) |
| Fear Test Subjective Fear Rating (0-8): mean (s.d.) | 3.87 (1.40) | 4.28 (1.15) |
| Affective Stroop Amygdala: mean (s.d.) Beta value | -0.05 (.18) | 0.07 (0.29) |
| Affective Stroop Reaction Time: mean (s.d.) seconds | 0.93 (0.16) | 0.96 (0.17) |

### Double-blinded placebo control and target pattern induction

Following neuro-reinforcement, participants were unable to correctly guess the identity of their neuro-reinforcement target (see *Supplemental Results* for more details), indicating they remained blind. Furthermore, the target phobic decoder showed a greater activation likelihood during neuro-reinforcement than the control phobic decoder (t(17)=12.63, p<0.001) (Fig. 2B, see *Supplemental Results* and *Supplemental Figure S2* for more detail), indicating successful nonconscious activation of the target representation during neuro-reinforcement.

### Amygdala Response (H1 and H5)

Before neuro-reinforcement, there was a significant amygdala response for both the target phobia (*t*(17)=2.20, *p*=0.042) and control phobia (*t*(17)=2.27, *p*=0.037) compared to neutral animals as confirmed by one-sample t-tests performed on the baselined parameter estimates. There was no difference in amygdala responses between the target and control phobias prior to neuro-reinforcement (*t*(17)=0.85, *p*=0.41). This indicates successful capturing of threat responding in the amygdala for phobic images.

Following neuro-reinforcement, there was a significant interaction between phobia type (target/control) and time (pre/post) shown by a 2 (condition) x 2 (time) repeated-measures ANOVA (*F*(1,15)=5.52, *p*=0.033, η_p_ ^2^=0.269, Fig. 4A). This result indicates a greater reduction in amygdala response to target phobic images than to control phobic images following neuro-reinforcement. After neuro-reinforcement, the decrease in amygdala response was significant for the target phobia (contrast of marginal means: *t*(25.75)=2.09, *p*=0.046, mean difference [s.e.] = 0.60 [0.29], 95% CI: 0.04-1.16) but not the control phobia (contrast of marginal means: *t*(25.75)=1.51, *p*=0.14, mean difference [s.e.] = 0.43 [0.29], 95% CI: -0.13-0.99). These findings support our preregistered hypothesis H1 that amygdala activation would be selectively reduced for the target phobia following neuro-reinforcement (see *Supplementary Results* and *Supplemental Figure S3*. for the H5 results). At post-test, there was no significant difference in amygdala response to the target and control phobias (contrast of marginal means: *t*(25.75)=0.112, *p*=0.91).

**Figure 4.**
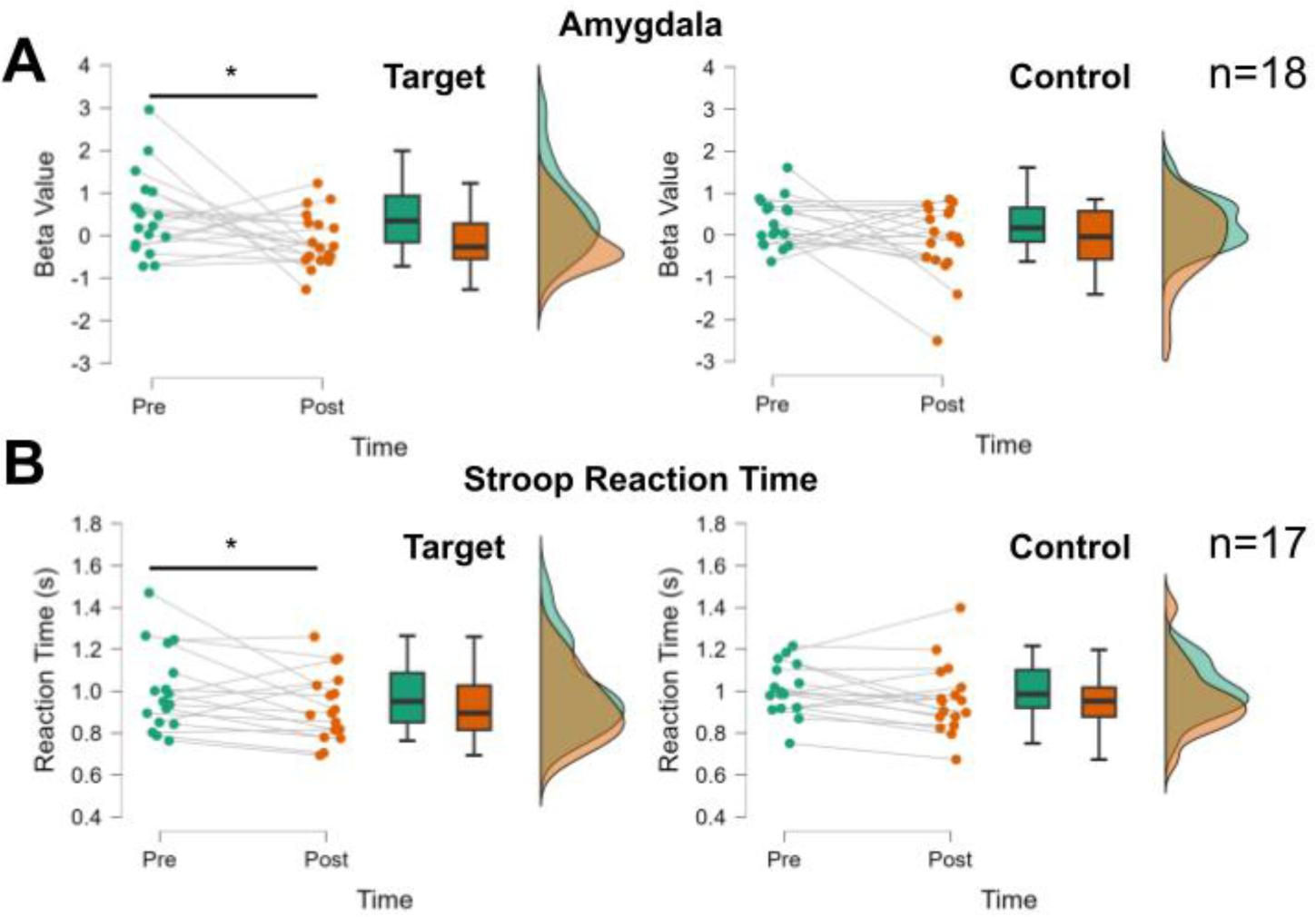
Changes in fear test amygdala responses and affective Stroop reaction times following neuro-reinforcement. (A) Amygdala response in the fear test showed a greater decrease in the Target than Control phobias following neuro-reinforcement. (B) Response times in the affective Stroop task showed a greater decrease in the Target than Control phobias following neuro-reinforcement. * *p*<0.05 indicate significant Time (pre/post) effect in the Target condition when controlling for days of neuro-reinforcement and number of phobias.

### Skin Conductance Response (H2)

Our findings did not support hypothesis H2. We did not detect a pre-treatment phobia response in SCR data in the 9 participants with complete SCR data, using one-sample t-tests on baseline-corrected SCR values for either target (*t*(8)=0.86,*p*=0.42) or control (*t*(8)=0.38,*p*=0.71) phobias. Given no significant pre-existing SCR response, no further statistical testing was performed.

### Self-Reported Fear (H3)

There was no significant change in self-reported fear levels for the 17 participants with complete behavioral data during the fear test in response to either the target phobia (*t*(16)=-1.52, *p*=0.15, mean difference [s.e.] = -0.24, 95% CI: -0.86-0.13) or the control phobia (*t*(16)=-0.56, *p*=0.58, mean difference [s.e.] = -0.08, 95% CI: -0.39-0.23), supporting our preregistered hypothesis H3. These findings match previous findings that self-reported fear levels in this task are not modulated by neuro-reinforcement (17).

### Affective Stroop (H4)

Results are reported from the 17 participants (5 participants in the 3-session group, 6 participants in other groups) with complete brain and behavioral data during the affective Stroop task. Before treatment, reaction times for phobic stimuli were significantly slower compared to responses to neutral stimuli (*t*(16)=2.64, *p*=0.018, d=0.64, mean difference [s.e.] = 0.067 [0.025], 95% CI: 0.017-0.12), confirming our preregistered hypothesis H4i. Slower reaction times for phobic stimuli indicate that attention is successfully captured by phobic stimuli in this task Importantly, there were no differences between reaction times to target and control phobic images pre-treatment (*t*(16)=0.91, *p*=0.38). Following neuro-reinforcement, there was a borderline significant interaction between phobia type (target/control) and time (pre/post) (*F*(1,14)=4.373, *p*=0.055, η_p_ ^2^=0.238), such that reaction times to the target phobia were faster following neuro-reinforcement than they were to the control phobia (Fig. 4B). Importantly, despite not quite reaching the significance threshold, the partial eta-squared effect size indicates a large effect. Specifically, there were significantly decreased reaction times to target phobia stimuli from pre-treatment to post-treatment (contrast of marginal means: *t*(18.13)=2.32, *p*=0.032, mean difference [s.e.] = 0.076 [0.033], 95%CI: 0.011-0.14) but not for control phobic stimuli (contrast of marginal means: *t*(18.13)=1.30), *p*=0.21, mean difference [s.e.] = 0.043, 95%CI: -0.022-0.11). Selectively decreased reaction times for the target phobia indicate that attention is captured less by the target phobia following neuro-reinforcement. At the post-test timepoint, there was no significant difference in reaction time to the target phobia compared to the control phobia (*t*(18.13)=-1.12, *p*=0.276). Reaction time effects by dosage group for H5 are reported in *Supplemental Figure S4.* Amygdala responding during affective Stroop (H4.iii) is reported in *Supplemental Results and Supplemental Figure S5A, S5B, S5C, and S5D*.

## Discussion

In a double-blind placebo-controlled clinical trial, we investigated whether multi-voxel neuro-reinforcement could nonconsciously intervene on specific phobia. In line with our hypotheses, we found evidence of large effects for specific reduction in amygdala reactivity (H1) and reduced attentional capture in an affective Stroop task (H4), though the interaction did not reach significance in this latter case. Importantly, these findings were obtained using surrogate participants to determine the target of neuro-reinforcement. Consequently, this study supports the ability of decoded neuro-reinforcement to be performed without exposing patients to the feared stimulus. Furthermore, our findings were obtained using a double-blind procedure, a level of rigor that is rarely achieved by other psychological interventions.

Decreases in amygdala responses and Stroop reaction times to phobic stimuli represent changes in physiological and reflexive responses to threat (21,33,34). These changes may represent ‘preconscious’ responses to feared stimuli due to their automatic and reflexive nature (35–41). Consistent with our hypotheses (H3) and prior findings (15), no effects were observed with respect to explicit subjective ratings of fear. As subjective fear ratings were not close to ceiling, this may be due to the task not eliciting large amounts of fear because participants are only viewing images of animals. Future studies should examine subjective fear to more ecologically valid in vivo exposures as a function of neuro-reinforcement. Alternatively, this pattern of results may suggest that implicit neuro-reinforcement is more effective for automatic physiological responses to threat compared to the subjective experience of fear itself. The discordance across response modalities would be consistent with a higher-order theory of emotion in which subjective mental experience operates via different mechanisms than physiological threat responses (21,33,34,42–45). While an effective treatment would ultimately aim to reduce subjective fear experiences when confronting phobic stimuli, neuro-reinforcement could represent an important first step prior to exposure therapy. For example, with reduced physiological threat responses, the reduction of subjective discomfort during traditional exposures may occur at an increased rate as subjective feelings come into alignment with already decreased physiological responding.

Furthermore, following neuro-reinforcement, results from the affective Stroop task showed statistical trends with large effects for reaction times decreasing for the target phobia relative to the control phobia (H4ii). In addition to providing further support for specific target engagement by neuro-reinforcement, this result suggests that individuals may be less reflexively avoidant of their phobia following neuro-reinforcement. If this is the case, patients may be more willing to persist in exposure that is conducted following neuro-reinforcement, leading to lower rates of attrition.

To test this hypothesis, future studies should complement neuro-reinforcement with a behavioral-approach task to investigate whether physiological symptoms are decreased when approaching the target phobia following neuro-reinforcement. If patients are more willing to approach the feared animal following neuro-reinforcement, then neuro-reinforcement may be a helpful complementary treatment alongside traditional exposure for ensuring the most comfortable treatment regimen possible.

Results of this experiment paired with our previous investigations (9,16,17) have collectively demonstrated the reduction of amygdala activity, implicit behavioral responses, and SCR using multi-voxel neuro-reinforcement, indicating extinction learning can occur nonconsciously. Exactly how this is accomplished remains an open question. The rationale for our methodology is based on exposure treatment with preliminary models supporting an exposure-like effect (9). The pattern of brain activity participants activate during neuro-reinforcement corresponds to a category-level visual representation of phobic animals. Our previous study also indicates that activation of this pattern alone does not generate an amygdala response (17), suggesting that the amygdala response profile is not altered itself during neuro-reinforcement. By inducing repeated activation of the visual representation, perhaps local connectivity changes impact how threat and fear systems respond to the natural presence of this representation during perception, achieving an extinction-like effect. Future studies should examine resting-state connectivity patterns before and after neuro-reinforcement to determine if neural changes are localized to visual or emotional processing areas.

The finding that something like extinction can occur nonconsciously using multi-voxel neuro-reinforcement is consistent with other studies using very brief exposure (VBE) (18,23,24). Our results theoretically support this paradigm while satisfactorily eliminating any doubt that some level of conscious awareness or threat processing is responsible for the observed effects in VBE. Future research should explore the overlap and differences between these strategies.

The current study is not without limitations however. As we did not detect SCR to phobic stimuli in our group of participants, we were unable to test one of our preregistered hypotheses (H2) that neuro-reinforcement would lead to reduced phobic SCR responding. This may have arisen from technical limitations, a large portion of participants being non-responders, or our relatively limited sample size. Similarly, the current study lacked the statistical power to test one of our other main hypotheses (H5); a between-subjects analysis of how much neuro-reinforcement is sufficient to achieve the desired outcomes. This limitation is directly due to our smaller-than-planned sample size (18 compared to 30 participants), a shortcoming that was due to the COVID-19 pandemic and funding body policies out of our control. This smaller sample size similarly limits the certainty that can be placed in our observed effect sizes - a limitation that should be addressed in a future study with a larger sample size. Moreover, the current study was limited by lack of a follow-up visit to determine how long effects of neuro-reinforcement may last. Future studies should explore how long neuro-reinforcement effects last following the intervention by re-testing participants weeks or months after neuro-reinforcement is completed.

Additionally, our primary measures, while important measures for neurocognitive understanding of fear and threat processing, are not traditionally treatment-targeted symptoms of specific phobia. In future studies, an independent session should be conducted following neuro-reinforcement examining real-life avoidance behaviors and fear levels. Future studies will be needed to understand how these observed changes translate to real-life avoidance and fear responses or perhaps using virtual reality as a first step.

While our use of a within-subjects placebo-control enabled us to test our intervention in a double-blind fashion, this design did not allow a between-subjects comparison with a placebo group. This should be addressed in a future study using a between-subjects design with treatment and placebo groups based on random assignment.

In summary, this study represents the first clinical trial of multi-voxel neuro-reinforcement for reducing fear and threat responses in specific phobia. This procedure demonstrated the ability to lessen physiological, reflexive responses to specific phobia through reduced amygdala activation as well as less attentional capture by phobic stimuli. These findings provide a promising foundation to attempt larger-scale replications in clinical cohorts. Through advents in virtual reality, these responses can also be investigated in future studies using more realistic and immersive stimuli (46–50). This nonconscious procedure produces minimal discomfort in patients with very low rates of attrition. Consequently, neuro-reinforcement may serve to complement current conventional psychotherapy approaches while providing a more tolerable experience for patients seeking treatment.

## Data Availability

All data produced in the present study are available upon reasonable request to the authors.

## Acknowledgments

The authors would like to thank study coordinators Ana Costello, Annelise Murillo, and Shawn Wang for their assistance to this study through participant recruitment and data collection. HL & MGC received financial support from the US National Institute of Mental Health (R61MH113772). HL received additional support from Templeton World Charity Foundation (RA537-01). MK was supported by AMED under grant number JP18dm0307008. VT-D received financial support from the Fonds de Recherche du Québec - Santé (FRQS) and the Fondation de l’Institut Universitaire en Santé Mentale de Montréal. This work was (partially) supported by Innovative Science and Technology

Initiative for Security Grant Number JPJ004596, ATLA, Japan.

## Author Contributions

Study design & conception – CAC, HL, MK, MGC, VTD. Data acquisition and analysis – CAC, VTD. Writing & Editing – CAC, HL, MK, MGC, VTD.

## Supporting Information

See Supplemental Information for Supplementary Methods and Results.

## Supplemental Information

### Supplemental Methods

#### Participants

##### Participant Recruitment

Recruitment was accomplished through flyers, campus website announcements, and posting on online forums (e.g. Nextdoor, etc). Participants completed the modified Fear Survey Schedule (1) in order to identify healthy controls who reported no phobias and individuals who endorsed at least two specific phobias of animals from the ones included in our image dataset. Participants were excluded if they did not meet criteria for MRI scanning safety.

##### Diagnostic Assessment

All participants underwent a diagnostic interview, using the Anxiety Disorders Interview Schedule-5 (2), administered by trained and reliability certified study staff (Bachelors degree), with each interview reviewed for final consensus by the Principal Investigator (MGC).

Participants were excluded if they 1) did not have normal/corrected to normal vision or hearing; 2) unable to understand informed consent or could not complete the consent form correctly; 3) unable to respond adequately to screening questions; 4) unable to maintain focus/stillness during assessment; 5) had a history of neurological disease or defect; 6) were diagnosed with PTSD, OCD, SUD, current MDD, Bipolar, Psychosis, or any other neurologic diagnoses or unstable serious medical conditions (all assessed using the ADIS-5); 7) currently prescribed psychotropic medication. Participants were not screened for active behavioral treatments.

##### Groups

*Healthy Control Group*: No animal type specific phobias or fears, ascertained from administration of the ADIS-5.

*Phobia Group*: Rated by a diagnostic interviewer (using the ADIS-5 (Brown et al., 2014) to have at least moderate fear or avoidance of at least two animals, with each one associated with an overall rating indicative of at least mild clinical severity. Fear and avoidance were each rated by the interviewer on a 0-8 point scale (0 = no fear/never avoids, 8=very severe fear/always avoids). Clinical severity was rated by the interviewer on a 0-8 point scale that combined symptom severity, distress and impairment associated with each animal stimulus (0=no symptoms, distress and impairment, 8=very severe symptoms, distress, and impairment). Phobias were only eligible if the clinical severity rating was at least mild (a score of 2+). For the 23 participants that were enrolled and started a pre-treatment session, participants had a mean (s.d.) of 2.39 (0.65) phobias. Target phobias had a mean (s.d.) fear rating of 5.17 ( 1.11), avoidance rating of 5.30 (1.43), and clinical severity rating of 4.65 (1.33). Control phobias had a mean (s.d.) fear rating of 5.91 (1.00), avoidance rating of 5.78 (1.24), and clinical severity rating of 4.70 (1.26).

#### MRI scanning parameters

All fMRI data were acquired on a 3T Siemens Prisma scanner using a 32-channel head coil at the UCLA Ahmanson-Lovelace Brain Mapping Center.

##### Decoder Construction

Across 6 task runs during decoder construction, fMRI data were collected with a multi-band sequence with an acceleration factor of 8 and phase encoding in the posterior (P) to anterior (A) direction in order to minimize dropout in the ventral temporal brain area. Voxel sizes were 2.0x2.0x2.0mm^3^ with a 208x208mm^2^ Field of View. Images were collected across 72 interleaved slices with a TR of 800ms, TE of 37.00 ms, and flip angle of 52 degrees. Anatomical data were collected using a T1-weighted imaging sequence with volumetric navigators (vNAV) with prospective motion correction (TR: 2500ms/TI: 1000ms/Flip Angle: 8.0 degrees/Voxel Size: 0.8x0.8x0.8mm/Matrix Size: 256x256/Num. Slices: 208/Slice Thickness: 0.8mm).

##### Multi-voxel neuro-reinforcement

Prior to the cessation of data collection due to the COVID-19 pandemic, fMRI data during the fear test task and affective Stroop task were collected across 2 runs each using the same sequence described for *Decoder Construction* for 7 participants. However, during the COVID-19 shutdown, this sequence was replaced with a similar but modified sequence better tailored for capturing BOLD activity in subcortical regions such as the amygdala. This replacement sequence used for the remaining 11 participants was a multi-band sequence with an acceleration factor of 6 and phase encoding in the A-P direction. Voxel sizes were 2.0x2.0x2.0mm^3^ with a 192x192mm^2^ Field of View. Images were collected across 72 interleaved slices with a TR of 1000ms, TE of 30.00ms and flip angle of 60 degrees. Accompanying Spin Echo Field Maps were collected in opposing phase encoding directions (A-P/P-A) before functional runs in order to be used for offline distortion correction. FMRI data during online neuro-reinforcement were collected using a multi-band sequence with an acceleration factor of 6 and phase encoding in the P-A direction to minimize dropout in the ventral temporal area. Additional parameters were voxel size: 2.0x2.0x2.0mm^3^, FOV: 208x208mm^2^, num. slices: 72, TR: 1000ms, TE: 37.00 ms, and flip angle: 60 degrees.

Importantly, this change in scanning sequence was only for the fear test task and affective Stroop task. No changes were made that could impact the building of decoders, or multi-voxel neuro-reinforcement itself. Additionally, this scanning sequence change had no effect on overall amygdala Beta estimates in the fear test task (*t*(16)=-0.391, *p*=0.70) or the affective Stroop task (*t*(15)=0.853). Hence, it is highly unlikely any findings in this study are due to the effects of this change in sequence during the covid shutdown.

#### Decoder Construction

##### Decoder Construction: task

In place of phobic images, phobic participants viewed happy human faces using stimuli from the Chicago Face Database and NimStim Set of Facial Expressions (3,4). These stimuli have their emotional expression verified by independent raters and were used to provide a non-disturbing stimulus replacement that was sufficiently orthogonal to the task image set of animals and objects. The decoder construction task consisted of 6 runs of 600 trials each. Each trial consisted of a .98 second image presentation with no inter-trial interval. This rapid event-related design was used to maximize the number of images each participant viewed. To ensure attention, participants were given the task of pressing a button each time the image category changed (i.e. a 1-back task). Image categories were presented in chunks of 2, 3, 4, or 6 consecutive images.

##### Decoder construction: fMRIprocessing

Decoder construction fMRI data were processed using a combination of SPM12 (Statistical Parametric Mapping; www.fil.ion.ucl.ac.uk/spm) and custom python scripts using pyMVPA and sklearn packages (5,6). All 6 runs of the task were concatenated and preprocessed in SPM using default parameters unless otherwise explicitly specified. Data were realigned to the first image from the first run of the task and segmented into tissue classes. Anatomical and functional data were coregistered using the gray matter image from segmentation as a reference. Motion was then regressed out of the functional data using the 6 head motion parameters from realignment. Single-trial estimates were then generated with pyMVPA using the least-squares 2 (LS-2) method (7) in which a separate GLM is computed for each trial where the current trial is assigned to one regressor while the remaining trials are equally split between two “rest” regressors.

Using hyperalignment, single-trial estimates from healthy controls in the target brain region (ventral temporal cortex) were functionally transformed to the current phobic participant’s brain and used to train a machine-learning pattern classifier (decoder) using the phobic images that the participant did not see (Fig. 1). To ensure double-blind treatment target selection, the target for treatment was automatically selected by a computer program that calculated which phobic category had the highest cross-validated area under the receiver operating characteristic curve (AUC) during binary one vs. all classification.

To determine AUC metrics, a 6-fold cross-validation (CV) procedure was used. FMRI data for each participant were loaded and masked to the ventral temporal (VT) area in their own native space using an anatomical mask derived from combining the entirety of the Freesurfer parcellations of the fusiform, lingual, parahippocampal, and inferior temporal areas (8). Single-trial parameter estimates were standardized by feature within subject and within each of the 6 task runs. The data were split into 6 folds for training and testing based on the 6 runs completed by each participant. That is, for each CV split, the withheld testing set consisted of all the data from each participant for one of the six task runs. The remaining preprocessing was calculated using only the training data to avoid overfitting. As hyperalignment requires a stable number of features across participants, 1000 voxels were selected within the VT area via F-test to select which voxels accounted for the most variance elicited by all image categories across all training trials. For each phobic participant, a unique set of hyperalignment transformation parameters into the common model space was calculated for the current phobic participant and all healthy controls. The fitting of the hyperalignment parameters was done using trials for all image categories except the current participant’s phobias. For example, if a phobic participant had spider and snake phobias, all spider and snake trials were withheld from all participants when fitting the transformation parameters.

After hyperalignment transformation parameters were determined, the data from all healthy controls were moved into the native space of the current phobic participant by transforming the data into the common model space and then reverse transforming the data from the common model space into the native space of the current participant. The transformed data included the previously withheld phobic category images from the healthy controls as well as the testing dataset.

With all data in the current participant’s native space, class sizes (target vs. non-target image categories) were balanced by random undersampling balanced between the 39 non-target image categories. Following previous work (9), a Sparse Multinomial Logistic Regression (SMLR) classifier was trained to perform binary (one-vs-rest) classification between the potential target category and all remaining categories (10). AUC scores for each CV split were calculated based on classifier estimates.

For the final decoder to be used in neuro-reinforcement, the same procedure was performed but trained using all 6 runs of data.

#### Pre/Post Test

##### Fear test: task

During each trial, a fixation cross was presented for 3-7 seconds, followed by a static image for 6 seconds. After the static image, a blank screen was displayed for 4-12 seconds followed by a prompt to enter how fearful they found the image on a 7-point scale. These ratings were used as the subjective fear ratings to test hypothesis H3. Images displayed either belonged to the target phobia, control phobia, neutral animal, or neutral object categories. Neutral animals and objects were randomly selected based on categories for which a given participant reported no fear during their diagnostic interview. Participants completed two runs of 15 images each with a self-paced break between runs. Within each run, they viewed 5 target phobia images, 5 control phobia images, and 2-3 neutral animal/object images, counterbalanced across runs. The first image of each run was a neutral object, always immediately followed by either a target phobia or control phobia image, counterbalanced across runs. The remaining images within a run were randomly selected from each category.

##### Fear test: fmri processing

FMRI task runs were distortion corrected using FSL’s topup (11,12) according to spin echo field map sequences collected in opposite phase-encoding directions. Due to technical issues with spin echo field map collection, 5 participants were excluded from distortion correction. Anatomical T1 images were brain extracted using bet (13). Then, preprocessing and ICA-decomposition were performed using FSL’s melodic and FEAT (FMRIB’s Software Library, www.fmrib.ox.ac.uk/fsl). During preprocessing, fMRI data were motion corrected using mcflirt (14), brain extracted using bet (13), spatially smoothed with a Gaussian kernel of FWHM 4.0mm, intensity normalized, and highpass filtered with a gaussian-weighted least-squares straight line fitting with sigma=50.0s. Images were then registered to the standard MNI space using FLIRT and then refined using nonlinear registration with FNIRT (14,15). Registration of multi-band images were improved by using a high-contrast single-band reference image collected at the start of each functional run as an initial reference image for registration.

ICA components were then manually investigated with components resulting from movement or other sources of noise removed. To further account for movement, data were processed with the Artifact Detection Tools (ART, https://www.nitrc.org/projects/artifact_detect) toolbox to generate motion regressors and identify outlier timepoints for censoring. First-level GLMs were then calculated in SPM12 with a temporal derivative to account for slice-timing differences. Regressors were fit for the onset of target phobia, control phobia, neutral animal, and neutral object images with a duration of 0 seconds to model the event-related response. Following previous work (9), only the first 2 trials within each run were analyzed for target phobia and control phobia images.

Bilateral amygdala masks were generated from the automatic Freesurfer segmentation of the T1 image and transformed into the participant’s native functional space. Average parameter estimates were extracted from the Amygdala using marsbar (16). Average parameter estimates for phobic stimuli were then corrected to baseline by subtracting the average amygdala response to the neutral animal from the target phobia and control phobia, within runs. Baseline-corrected phobia responses were then averaged across runs for pre-treatment and post-treatment sessions.

##### Affective Stroop: task

The task started with a 1 second red fixation cross and then a brief (300 ms) image from either a phobic or neutral control category. As soon as the image appeared, participants were instructed to, as quickly and accurately as they could, make a size judgment about whether the presented animal could fit in their hand (i.e. is it the size of your hand or smaller?), by pressing one of two buttons with their index and middle finger to indicate yes or no. Response-key mappings were counterbalanced across participants. There was a 1.2 second response period (indicated by a blue fixation cross) following stimulus offset for response entry followed by a fixed 1 second inter-trial interval. Stimuli were selected from 7 animal categories: target phobia, control phobia, and 5 neutral animal categories. Similar to the fear test, neutral animal categories were selected from categories for which a given phobia participant reported no fear during their diagnostic interview. The task consisted of 210 randomly distributed trials split over 2 fMRI runs with a self-paced break between runs.

##### Affective Stroop: analysis

Reaction times were extracted for target phobia, control phobia, and neutral animal stimuli using custom scripts in MATLAB (Mathworks Inc., Natick, MA). Responses were coded as correct or incorrect based on unanimous agreement from 8 independent raters who judged whether each of the 30 potential animal categories was the size of their hand or smaller: unanimity was obtained for 24 animal categories; animal categories without consensus (bird, bat, fish, gecko, turtle, and guinea pig) were treated as correct as long as a response was recorded.

#### Multi-voxel neuro-reinforcement

##### Online real-time fMRI processing

Real-time fMRI processing for multi-voxel neuro-reinforcement was conducted in MATLAB with the decoded neurofeedback software developed at Advanced Telecommunications Research Institute International (https://bicr.atr.jp/decnefpro/software/). Incoming dicom images exported from the scanner were converted to nifti, realigned to a template image, the first dicom from the first run of the decoder construction task, then detrended based on all the data collected up to that TR in a given run. Proper alignment between the real-time fMRI data and the decoder construction data (on which the decoder was based) was ensured by correlating multi-voxel patterns between the real-time data and the decoder construction template. If pattern correlation fell below a threshold of 0.70 on a given trial, visual feedback was not displayed to the participant.

##### Monetary Reward

The size of the feedback disc determined the amount of reward the participant received at the end of each run, with their average feedback score determining the percentage of that run’s total bonus received. For example, an average feedback score of 60% resulted in 60% of the potential $6.00 bonus being received (i.e. $3.60). An additional bonus was also given when participants were able to generate a feedback score of 70% or more for 3 trials in a row. Participants were given an additional $2.00 per high-score streak bonus which was visually indicated by the feedback disc turning blue with a written message alerting them to their high-score streak.

#### Skin Conductance Response (SCR)

##### Data collection

Skin Conductance Response (SCR) was recorded in Acknowledge software via Biopac MP-150 system using the EDA-100C module and Ag/AgCl electrodes placed distally on the index and middle fingers of the left hand. SCR recordings were taken during pre-treatment and post-treatment MRI scanning sessions. Of the 18 participants analyzed in our main analyses, 5 participants had technical issues during data collection and 4 participants were non-responders showing no discernable SCR. Consequently, 9 participants were analyzed for SCR.

##### Data analysis

SCR recordings were analyzed with custom code in python utilizing the bioread package. SCR data were filtered with a 1st-order 5 Hz low-pass Butterworth filter to account for influences of the magnetic field in the MRI environment. SCR recordings were then epoched according to stimulus onset times during the Fear Test task from 2 seconds preceding stimulus onset to 5 seconds following stimulus onset. Epoch timecourses were baseline corrected according to the average activity during the 2 seconds before stimulus onset. Peak SCR values were then extracted for each trial epoch by taking the maximum SCR value in the time period of 1 second to 5 seconds following stimulus onset. If the peak SCR value was less than 0.02 microsiemens then it was coded as 0 following previous research (ref). Peak SCR values were then square root transformed in preparation for statistical analysis.

##### Self Report Questionnaires

The following self report questionnaires were administered at pre-treatment and post-treatment:

Depression, Anxiety, and Stress Scale (DASS-21) (17), Behavioral Inhibition/Behavioral Activation Scale (BIS/BAS) (18), Sheehan Disability Scale (SDS) (19), and Modified Fear Survey Schedule (1).

#### Supplemental Results

##### Hyperalignment decoding

Binary classification performance of hyperaligned decoders was estimated in a 6-fold cross-validation on decoder construction task data. Average target decoder AUC was 0.63 (0.03), which was significantly greater than the chance level of 0.50 (*t*(22)=20.3, *p*<0.001). This indicates that category-level visual representations can be significantly decoded in the brains of participants with specific phobia based on hyperaligned surrogate brain data from healthy controls.

##### Double-blinded placebo control

After neuro-reinforcement, the experimenter revealed to participants that neuro-reinforcement feedback had been based on the visual representation of one of their phobias. When asked to pick between two of their phobias (the target and control phobias, blinded to the experimenter), participants were unable to correctly guess the identity of their neuro-reinforcement target (43% accuracy; chance level 50%). Participants reported strategies for neuro-reinforcement that were unrelated to the target and control animal categories. Collectively, this indicates neuro-reinforcement was carried out in a double-blind fashion at an implicit level with participants unaware of the target of the intervention.

##### Target pattern induction

To assess the degree to which the desired pattern associated with the target phobic category was activated by patients during neuro-reinforcement, the feedback scores patients saw (representing degree of desired neural pattern activation) during neuro-reinforcement were compared to the scores patients would have seen if feedback had been based on the control phobic category pattern instead. Both these target and control scores were generated using the real-time pipeline - the only exception being the fMRI data was detrended across the entire run (versus how much data had been collected up to a given ‘trial’) as this is how the data were saved at the end of the neurofeedback program once the entire run had been collected. In the 18 participants analyzed for our primary outcome, the feedback was significantly higher for the target phobic category compared to what it would have been for the control phobic category (t(17)=12.63, p<0.001) (Fig. 2B). This result indicates that the desired target pattern was successfully activated by patients during neuro-reinforcement. Results for each individual day are reported in Supplemental Fig. S1.

##### Amygdala response during Stroop task

Amygdala responding during the affective Stroop task did not demonstrate the same interaction we observed during the fear test task (*F*(1,14)=1.075, *p*=0.317) counter to our pre-registered hypothesis H4iii. Additionally, in the affective Stroop task, a phobia response was not observed in response to the target phobia pre-treatment as tested with a one-sample t-test on the baselined parameter estimates (*t*(16)=0.19, *p*=0.85). This lack of significant phobia response pre-treatment could be due to the increased cognitive load of this task which required rapid, reflexive judgments as soon as the stimulus appeared (compared to fear ratings in the fear test which were input many seconds after the original stimulus disappeared). Additionally or alternatively, the amygdala may have habituated during the affective Stroop task as it was always immediately preceded by the fear test.

##### Between-subjects analysis of dosage effects (H5)

Although circumstances outside of our control (detailed in methods) prevented us from collecting a sufficient sample size to analyze the between-subject effect of dosage with sufficient power as we initially pre-registered, we report the pre-registered analysis here. When dosage (1, 3, or 5 days of neuro-reinforcement) is treated as a between-subjects factor in a 3 (between-subjects dosage: 1, 3, or 5 days of neuro-reinforcement) x 2 (within-subjects condition: target, control phobia) x 2 (within-subjects time: pre-treatment, post-treatment) repeated-measures ANOVA, we fail to find evidence in support of H5. The 3-way interaction between dosage, condition, and time is trending but not significant (*F*(2,14) = 3.236, *p*=0.07, η_p_^2^=0.316). Within each group, significant differences could not be detected for target responding pre-to post-treatment. The greatest effect detected was for the 3-day group for a reduction in amygdala responding to the target phobia pre-to post-treatment (*t*(5)=0.137, *p*=0.137). This lack of evidence in support of our pre-registered hypothesis H5 is most likely due to insufficient power (6 participants in each dosage group) to detect such between-subjects effect in the current design. Future studies will be needed to address the question of how the number of neuro-reinforcement sessions an individual receives affects reduced amygdala responses to feared stimuli.

##### Self-Report Questionnaires

A paired sample t-test for Depression, Anxiety and Stress Anxiety Subscale was marginally significant, t(17) = 2.06, p = .055: pre-test (M = 8.9, SD = 2.7) and post-test (M = 8.2, SD = 1.6) indicating a marginal decrease in anxiety following neuro-reinforcement. There were no effects for the depression subscale or stress subscale or the total DASS score.

##### Assessment of additional covariate

We could not anticipate how much variance would be present in the number of phobias amongst participants with multiple phobias. While the final analyzed sample had a mean (s.d.) number of phobias of 2.39 (0.65), some participants during recruitment had as many as 5-7 phobias. This not only indicates a more widespread experience of clinical fear but also introduces a potential difference during the decoder construction process as the total number of phobias had to be withheld from data preprocessing during the hyperalignment process. For these reasons, we elected to include the number of phobias as a covariate in statistical analyses. The interpretation of the reported results remains the same when this covariate is not included in the model. More specifically, the interaction effect for H1 was still significant ((*F*(1,16)=5.17, *p*=0.037, η_p_ ^2^=0.244) and the interaction effect for H4 which was near significance, remained non-significant (*F*(1,15)=0.041, *p*=0.84, η_p_ ^2^=0.000069).

### Supplemental Tables

**Supplementary Table 1.** Participant demographics by dosage group.

| **Race** | **1 day** | **3 day** | **5 day** |
| --- | --- | --- | --- |
| White | 2 | 3 | 4 |
| Black | 0 | 1 | 1 |
| Asian/Pacific Islander | 4 | 4 | 2 |
| Other | 1 | 0 | 0 |
| Not Reported | 1 | 1 | 1 |
| **Ethnicity** |  |  |  |
| Hispanic | 1 | 1 | 3 |
| Non-Hispanic | 7 | 6 | 5 |

DECNEF INTERVENTION FOR SPECIFIC PHOBIAS
| **Gender** |  |  |  |
| --- | --- | --- | --- |
| Male | 2 | 0 | 3 |
| Female | 6 | 7 | 5 |
| Non-binary | 0 | 0 | 0 |
| **Age: mean(sd)** | 25.5 (6.6) | 26.3 (12.9) | 27.6 (9.4) |
| **Education Level** |  |  |  |
| High School | 1 | 1 | 2 |
| Some College | 1 | 1 | 1 |
| Associates/2-year degree or higher | 6 | 5 | 5 |

### Supplemental Figures

**Supplemental Figure S1.**
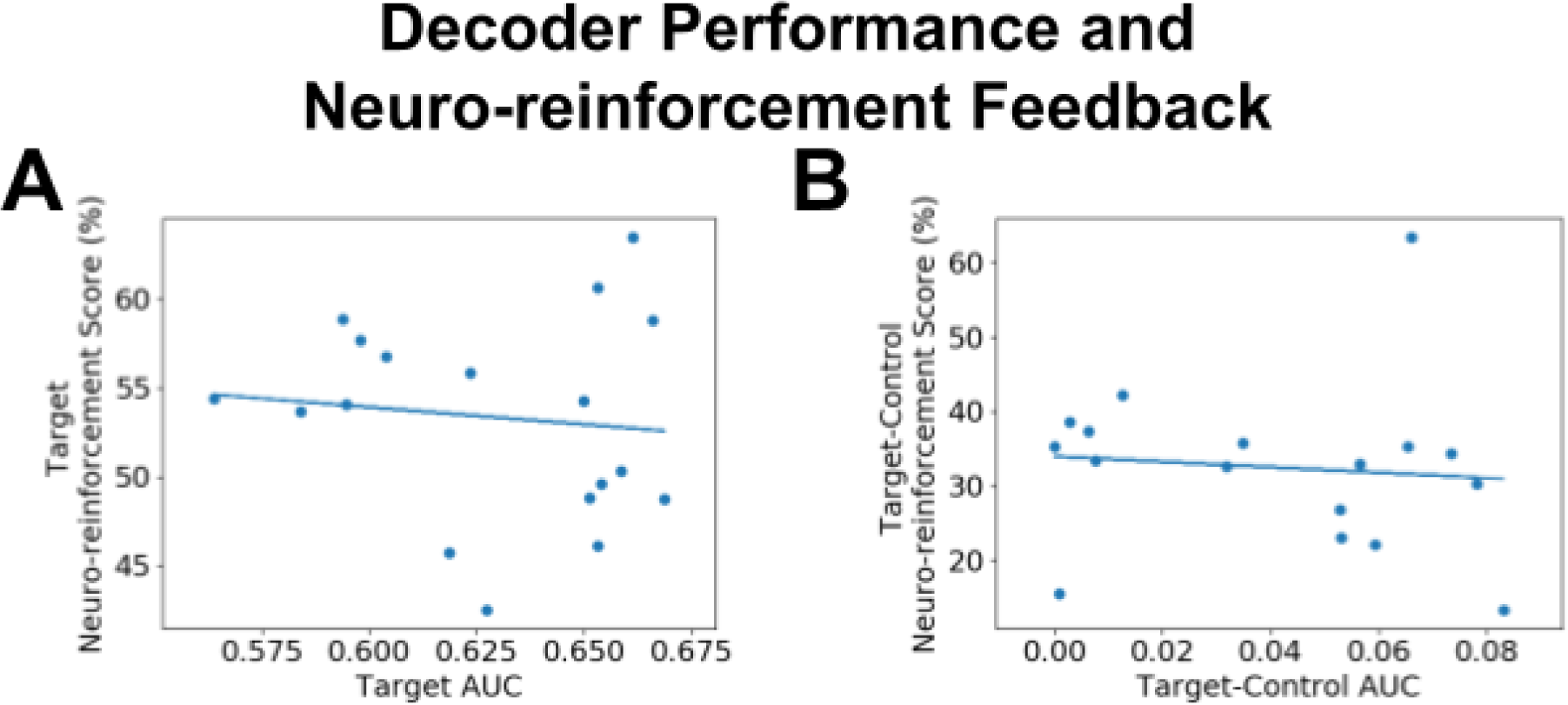
No relation between decoder construction performance and neuro-reinforcement scores. Scatter plots of average cross-validated AUC scores during decoder construction and neuro-reinforcement scores from neuro-reinforcement sessions. Solid lines indicate line of best fit. (A) Association for the target phobic category (B) Association for the difference between the target and control phobic categories.

**Supplemental Figure S2.**
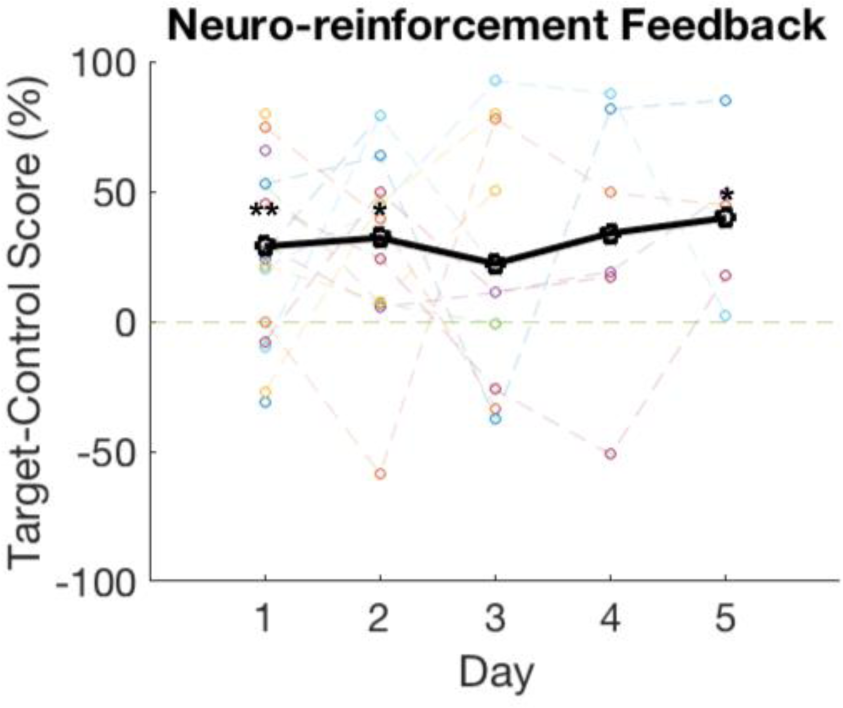
Neuro-reinforcement feedback for target versus control phobias by day. Daily averages of target minus control scores for each participant are plotted in colored data points. Each color codes for an individual participant across days, connected by dashed lines. Black data points represent average from all participants on a given day, connected via solid black line. Individual days with significantly greater target feedback compared to control feedback according to paired t-tests are marked with an asterisk. * *p*<0.05, ** *p*<0.01

**Supplemental Figure S3.**
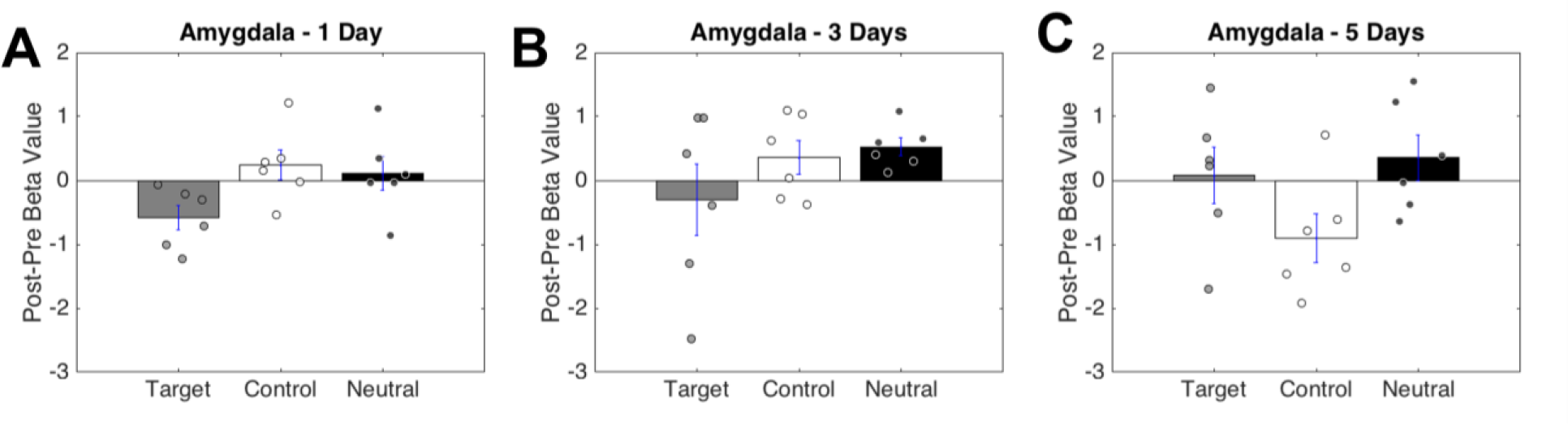
Amygdala response following neuro-reinforcement by number of days of neuro-reinforcement received. Panels show changes in responses to target phobia, control phobia, and neutral animal images from pre-neuro-reinforcement to post-neuro-reinforcement, quantified as post minus pre difference. Results for participants that received 1 day (A), 3 days (B), or 5 days (C) of neuro-reinforcement.

**Supplemental Figure S4.**
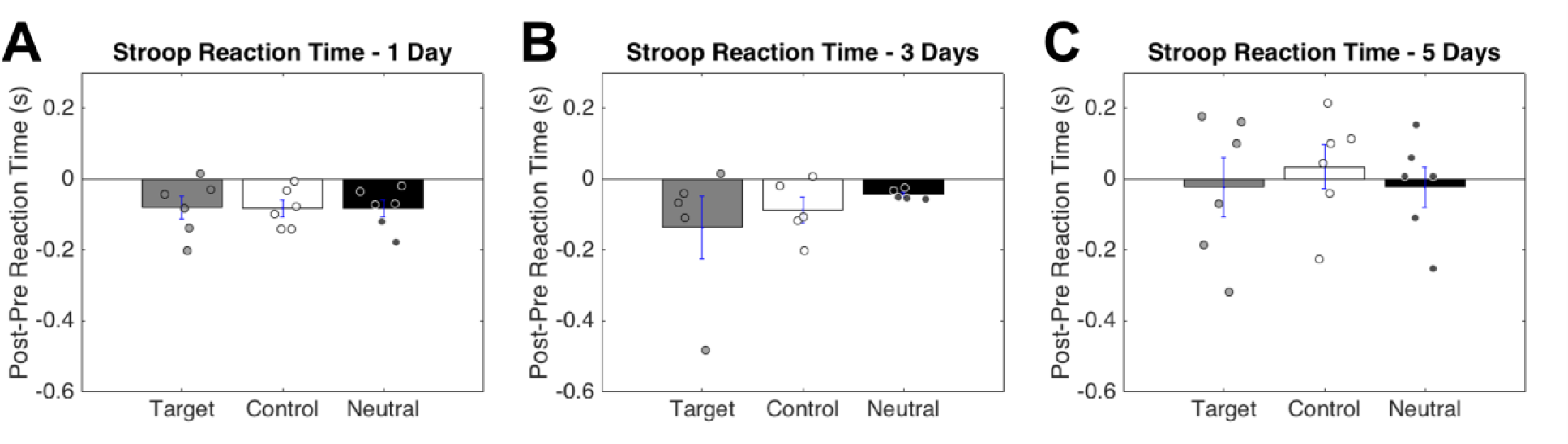
Reaction times in affective Stroop task following neuro-reinforcement by number of days of neuro-reinforcement received. Panels show changes in reaction times to target phobia, control phobia, and neutral animal images from pre-neuro-reinforcement to post-neuro-reinforcement, quantified as post minus pre difference. Results for participants that received 1 day (A), 3 days (B), or 5 days (C) of neuro-reinforcement.

**Supplemental Figure S5.**
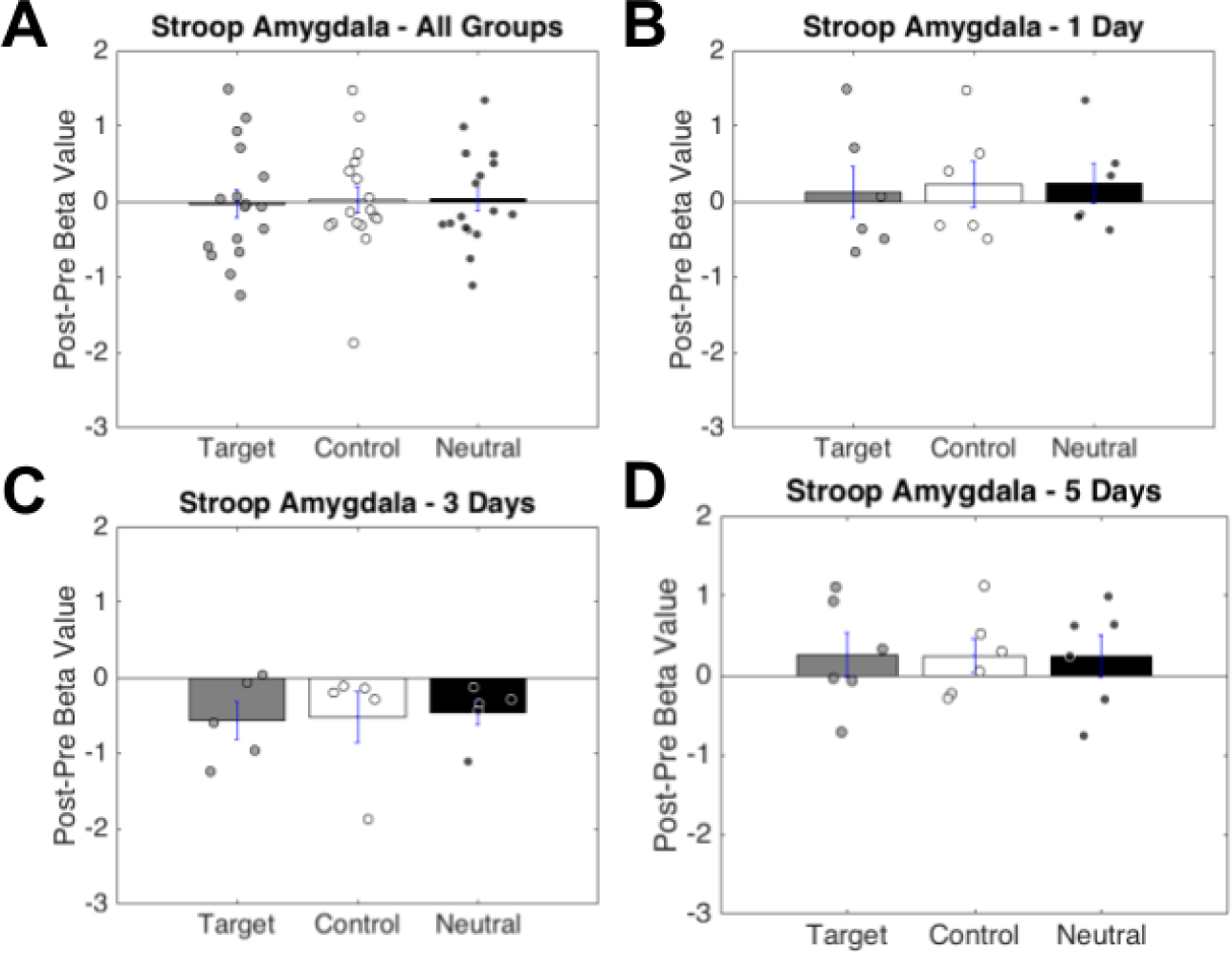
Amygdala response following neuro-reinforcement in affective Stroop task. Results for all dosage groups combined (A) showing post-treatment minus pre-treatment amygdala responses to target phobia, control phobia, and neutral animals in the affective Stroop task. Also, the 1 day (B), 3 days (C), and 5 days (D) of neuro-reinforcement are also shown for illustrative purposes.

